# Inverse proportionality between height and duration of epidemic peaks not observed for the COVID-19 epidemic in Japan

**DOI:** 10.1101/2022.03.23.22272840

**Authors:** Toshihisa Tomie

**Affiliations:** Changchun University of Science and Technology, Changchun, Jilin, China

## Abstract

The height of the epidemic peaks varied ten-fold, but the duration was almost constant independent of the peak height in the six times COVID-19 epidemics in Japan over the past two years. The observed relation between the peak height and duration contradicts the inverse proportionality, which is the essential conclusion derived from mathematical models for infectious diseases. We found that the peak height was inversely proportional to the number of rhinovirus patients. The literature has revealed the mechanism behind our found power of rhinovirus suppressing COVID-19. We discuss that the critical flaw of current mathematical models originates in the absence of the 0th power term of the number of infected people in the Kermack and McKendrick equation.

## 1. Introduction

Mathematical models are a powerful tool for understanding the epidemic situation of infectious diseases. When the population and microorganisms increase exponentially, the increase rate in the number of individuals decreases as the number approaches the environmental acceptance, and it converges to a steady value. This phenomenon follows the logistic equation (Ref. 1),

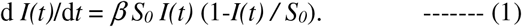

In the case of infectious disease, it is no longer the source of infection once the infected person is recovered or quarantined. In 1927, Kermack and McKendrick (Ref. 2) made the correction incorporating the recovery of the infected individuals,

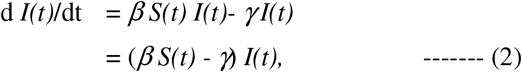

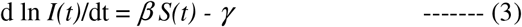

Here, *I(t)* is the cumulative number of infected individuals, *S(t)* is the number of susceptible persons at time *t, β* is the infectivity, *S*_*0*_ is the number of members of the society, and *γ* is the recovery rate of the infected individuals. We call this historic model the K-M model and Eq (2) the K-M equation.

Equation (3) tells that the infection does not start to spread if *β S*_*0*_ *-γ* <0. Contagious power *β* is proportional to the frequency and closeness of human contacts. Governments issue lockdown of cities to reduce *β*. The purpose of hand-washing and wearing a mask is to reduce the contagious power *β*. Vaccination prevents individuals from infection. But the principal purpose is to achieve herd immunity by reducing the number of susceptible people *S*_*0*_ for *β S*_*0*_ *-γ* <0. All measures for controlling infectious disease originate in the model.

We forecasted the end of the epidemic in Wuhan in 2020 February by Gaussian fitting of the change in the number of infected people, and the epidemic ended as forecasted (Ref. 3). Gaussian fitting originates in the fact that both the logistic function and the numerical solution of the K-M equation can be approximated by a Gaussian, *I (t)* = *I*_*0*_ exp (-((*t*-*t*_*0*_) / tau) ^ 2), near the peak of the epidemic. As is clear from Eq. (3), at the onset and end of the epidemic, the number of infected people increases and decreases exponentially, so the deviation from a Gaussian is large. The epidemic profile of influenza in the 2019/20 season in Japan is reproduced better by the numerical solution of the K-M equation than by a Gaussian (Ref. 4).

However, we noticed some things that mathematical models do not reproduce (Ref. 4). One is that the number of patients in actual diseases is far smaller than the model calculation. The ratio of the final integrated number of infected people to the number of populations is called the final size, *p (∞)*, which is given by (Ref. 4),

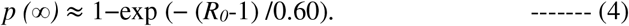

Here, *R*_*0*_ *= βS*_*0*_ */γ* is the basic reproduction number. For *R*_*0*_>1.5, *p (∞)*> 0.5.

Influenza in the 2019/20 season in Japan was reproduced by *τ*_*trans*_= 0.52 weeks and *τ*_*inf*_ = 1 week, where *τ*_*trans*_=1/*β S*_*0*_ and *τ*_*inf*_ = 1*/γ* (Ref. 4). Therefore, *R*_*0*_ =1.92, then, *p (∞)* = 0.78. However, the number of influenza patients in Japan is only about 10% of the national population each year. We cannot claim that the real number of influenza-infected people is eight times the number of people who consulted doctors. Many papers reported that the positive rate of PCR tests for influenza is about 40 %. (Refs. 5∼7). This rate is for people developing influenza-like illnesses. The rate should be lower for asymptotic people. The final size for influenza as large as 78% can never be possible. Definitely, there is a flaw in the model.

The other is that influenza patients are constantly detected, which is not describable in current mathematical models. This problem is not trivial either.

There is another and crucial problem with current mathematical models; the relationship between epidemic peak height and duration. With high infectivity, the number of infected people surges and peaks, infecting a significant proportion of the population, and the epidemic ends in a short period. If the infectivity is low, the number of infected people increases slowly, the peak height is low, and the epidemic extends over a long period before a considerable proportion of the population is infected. Hence, peak height and duration period are roughly inversely proportional. In actual infectious disease epidemics, however, the inverse proportionality between the height and width of the peak is not observed. This flaw is critical.

This paper discusses in detail the inverse proportionality between height and duration of epidemic peaks derived from the K-M model. We examine the relationship between the height and width of the COVID-19 epidemics in Japan. In six COVID-19 epidemics, peak height varied by one order of magnitude, but as with other infectious diseases, the epidemic duration was not inversely proportional and was independent of the peak height, which contradicts the essential conclusion from mathematical models. We found that the height of the six epidemic peaks of COVID-19 was inversely proportional to the number of rhinovirus patients, which agrees with the literature. We discuss that the critical flaw of mathematical models originates in the absence of the 0th power term of the number of infected people, *I(t)*, in the Kermack and McKendrick equation.

## 2. Analyzed data

All clinical data analyzed in the present paper are publicly available.

The data of positive people in Tokyo are from the Tokyo Metropolitan Government Corona Virus Infection Control Site. The positive rate is from the file in “Monitoring item (4) Positive rate of test” (Ref. 8). We calculated the age distribution using the data in “Attributes of positive people” (Ref.9).

The data on rhinovirus is from the file in the “Archive of Other Viruses” (Ref. 10) at the site of Infectious Agents Surveillance Report (IASR) at the National Institute of Infectious Disease.

## 3. Results

### 3-1. inverse proportionality between height and duration of epidemic peaks

Figure 1 shows the numerical integration of Eq. (2) for several values of *βS*_*0*_ in 1/days with the fixed *γ* of 1/ 3.1 days. For a high infectious power *βS*_*0*_, the number of infected people rises quickly and produces a high epidemic peak. The profile near the peak can be approximated by a Gaussian displayed by solid curves of parameters shown in the figure. The deviation of the fitting Gaussian from the detailed change calculated by the K-M model is not a problem in characterizing the epidemic because statistic fluctuation due to the small number of patients can be larger than the deviation at the skirts of the epidemic.

**Fig. 1:**
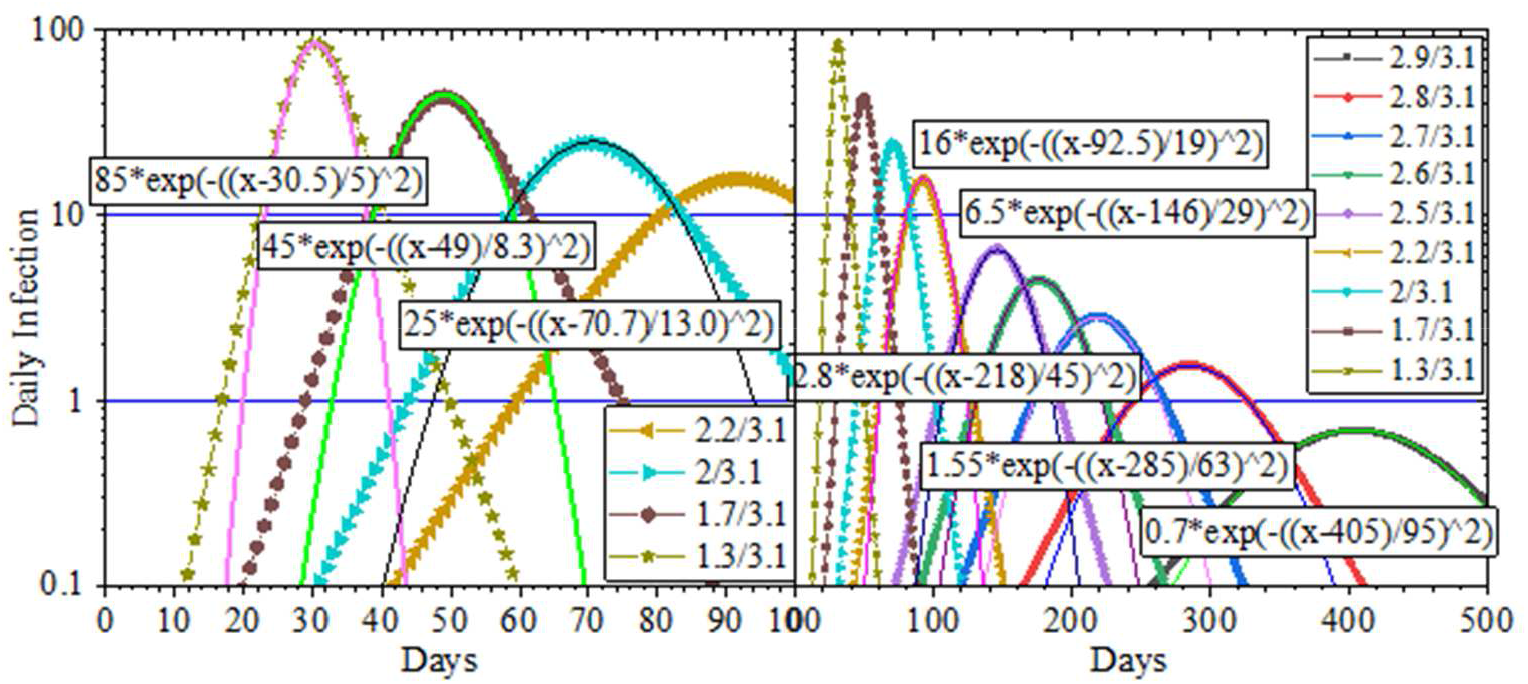
The numerical integration of Eq. (2) for several values of *βS*_*0*_ in 1/days with the fixed *γ* of 1/3.1 days. The profiles near the peak are fitted well by Gaussians of parameters shown in the figure.

Figure 2 summarizes the peak height and time constant of the fitting Gaussians, shown in Fig.1, as a function of *βS*_*0*_ – *γ*. The peak height (black squares) increases by 1.5 power of *βS*_*0*_ – *γ*. The time constant (red circles) is about 20 days when *βS*_*0*_ – *γ* is 1/ (10 days) and shortens in inverse proportion to *βS*_*0*_ – *γ*.

**Fig. 2:**
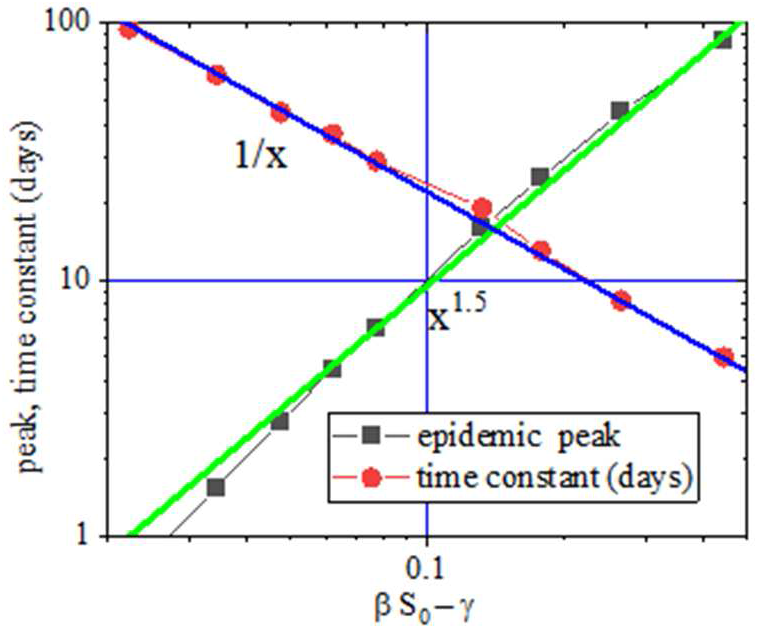
Summary of parameters of Gaussians in Fig.1. The height increases by 1.5 power, and the time constant shortens in inverse proportion to *βS*_*0*_ *– γ*.

### 3-2. the COVID-19 epidemic in Japan

We examine whether the essential result of mathematical models, the inverse proportionality of height and duration of epidemic peaks, is observed in the COVID-19. COVID-19 has been prevalent six times in Japan since the first epidemic in the spring of 2020. Figure 3 shows changes in the positive rate and the number of positives in public PCR tests in Tokyo (Ref. 8). The number of positives (black points) for the first epidemic peaked at 100 per day, followed by over 300 per day in the summer epidemic of 2020 and then followed by 1,000 for the winter, spring, and summer epidemic of 2021. Quite strangely, there were more than 200 positives in the valleys, exceeding the peak of the first epidemic. However, the positive rate (red dots) for the first epidemic was 30% higher than the valley values, about the same as the summer of 2021, and higher than the other three peaks. The low number of positives in the first epidemic was probably due to the low number of tests. As the number of tests increases, so does the number of positives. The positive rate is less affected by human factors such as the number of tests, and we better discuss the epidemic using the positive rate as claimed in Reference 11. In the present paper, we discuss epidemics by the positive rate, not by the number of positives.

**Fig. 3.**
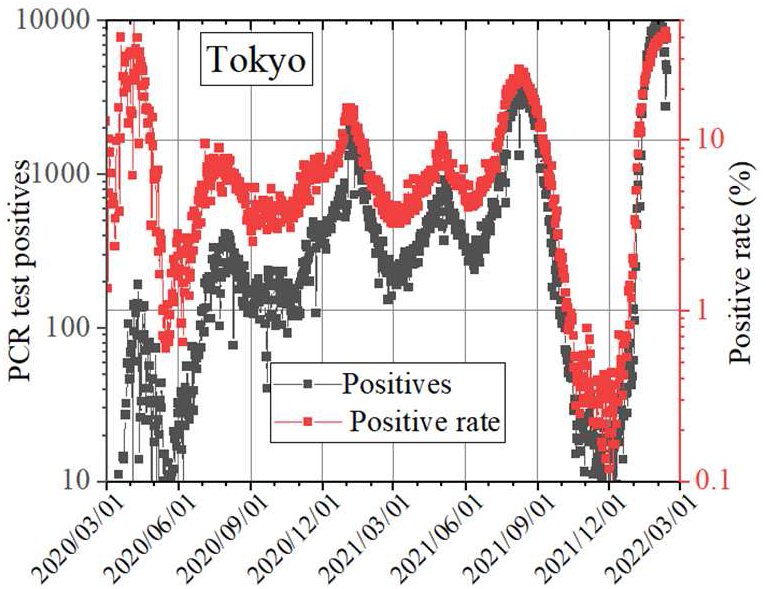
The changes of PCR test positives and the positive rate in Tokyo for the last two years. There have been six COVID-19 epidemics.

Peak values of the positive rate varied from epidemic to epidemic at 30%, 8%, 15%, 10%, 30%, and 40%. In the epidemic valleys in June and October 2020 and March and June in 2021, the positive rates were 1%, 5%, 5%, and 5%. But in October and November 2021, the positive rate was as low as 0.3%. Because of the large variation of the peak size, the COVID-19 epidemic in Japan is the optimal example for examining the validity of the present mathematical models.

### 3-3. Gaussian fitting of the COVID-19 epidemic

We take a close look at the variation of the epidemic of COVID-19. Infectivity *β* depends on the activity of the virus and the immunity of the person, and it can be seasonal. Therefore, we compare the epidemics in 2020 and 2021 by season.

Figure 4 shows the epidemics in spring (5th to 25th week). The COVID-19 epidemic can be approximated by a Gaussian of 0.03 + 0.28 * exp (-((x-14.5) /2.4)^2) in 2020, and 0.045 + 0.040 * exp (-((x-18)) in 2020.) /2.4) ^ 2) in 2021. The peak sizes were seven times different, but the time constants were the same of 2.4 weeks.

**Fig. 4.**
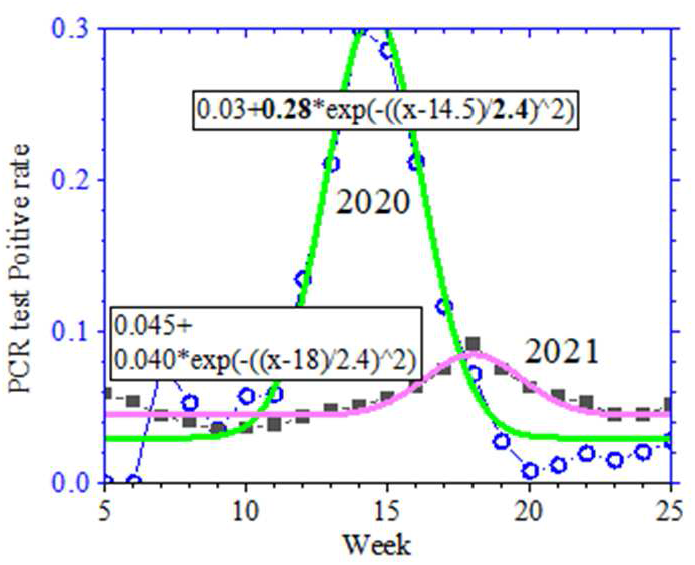
The spring epidemics in 2020 and 2021. The peak sizes were seven times different, but the time constants were the same.

The summer epidemics in Figure 5 can be approximated by 0.02 + 0.05 * exp (-((x-30.5) /4.5) ^ 2) in 2020 and 0.04 + 0.2 * exp (-((x-32.3) /3.7) ^ 2) in 2021. The size of the epidemics is four times different, but the time constant of Gaussians was about the same again, about 1.5 times that of the spring epidemic.

**Fig. 5.**
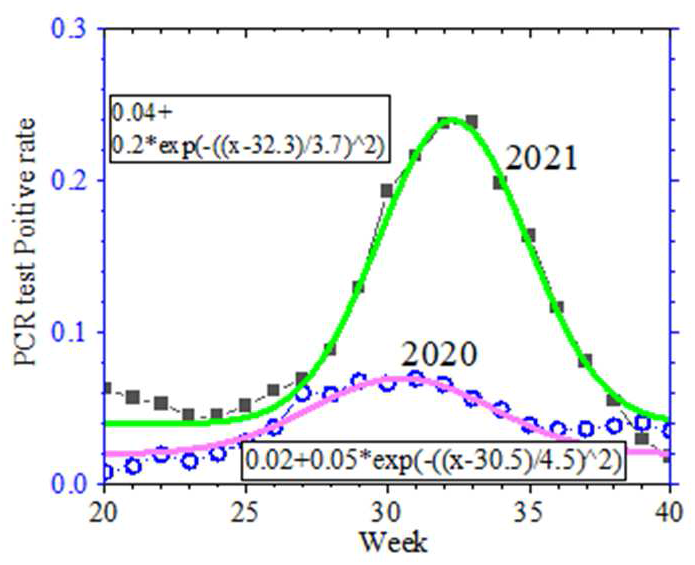
The summer epidemics in 2020 and 2021. The size of the epidemics is four times different, but the time constant of Gaussians was about the same.

The winter epidemics shown in Figure 6 can be approximated by 0.065 + 0.085 * exp (-((x-53.6) /1.7) ^ 2) in 2021 and 0.43 * exp (-((x-58.5) /4.3) ^ 2) in 2022. The time constant of the smaller epidemic in 2021 was as small as 1.7 weeks, and it was as large as 4.3 weeks for the larger epidemic in 2022.

**Fig. 6.**
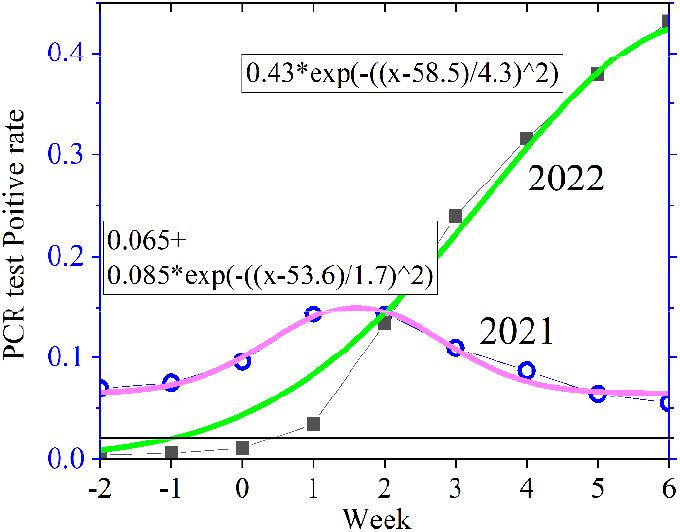
The winter epidemics in 2020 and 2021. The time constant of the weaker epidemic in 2021 was as small as 1.7 weeks, and it was as large as 4.3 weeks for the larger epidemic in 2022.

Figure 7 (a) summarizes the peak positive rate and the time constant of fitting Gaussians in Figs. 4 to 6. Time constant depended on the season and was independent of the peak height of the epidemics when the peak height varied by one order of magnitude.

**Fig. 7.**
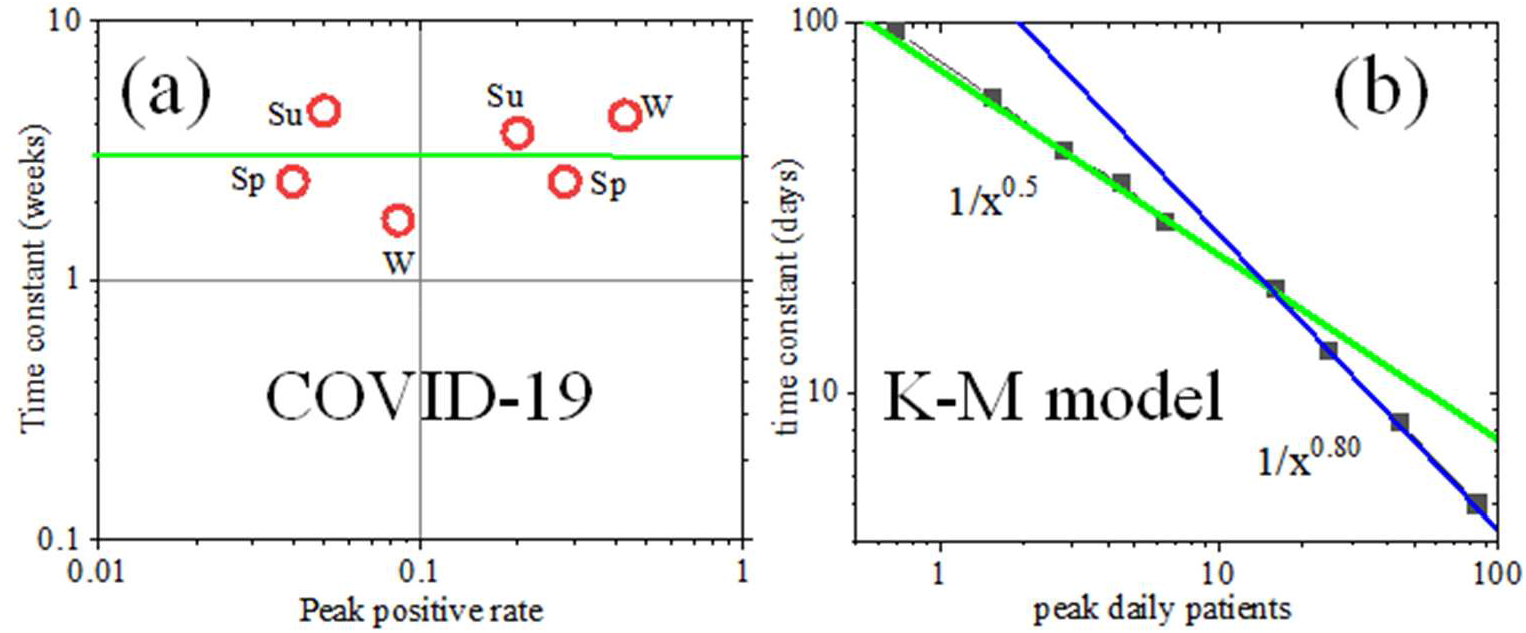
(a) Summary of the time constants and peak heights of fitting Gaussians shown in Figures 4 to 6. The “Sp”, “Su”, and “W” stand for spring, summer, and winter, respectively. The time constant was independent of the peak height. (b) Redrawing of Fig.2; the calculation by the K-M model. The time constant is shorter for a higher peak.

Figure 7 (b), which is redrawing of Fig.2, shows the time constant as a function of the peak daily patients calculated by the mathematical model. When the peak value is 15 or less, the time constant is shorter in inverse proportion to the square root of the epidemic peak, and when the peak value is 15 or larger, it shortens in inverse proportion to the 0.8th power of the peak height. Figure 1 shows the results of calculations with *γ*=1/3.1 days. Then, *βS*_*0*_ – *γ*=0.1 means *R*_*0*_-1 = 0. 31 and we get *p (∞)* ≈ 0.4 from eq. (4). For *R*_*0*_-1> 0.2, the value of exp (-(*R*_*0*_-1) /0.60) begins to saturate and the final size saturates. These calculations explain the slope change seen in Figure 7 (b). Figure 7 (a) for COVID-19 contradicts Figure 7 (b) from the K-M model.

### 3-4. seasonality of the COVID-19 epidemic

The time constant is independent of the peak height but is seasonal. The legends of Su, Sp, and W in Fig. 7 (a) stand for summer, spring, and winter, respectively. The time constant was larger in warmer seasons except for the “W” point at the highest positive rate.

The seasonality of the COVID-19 epidemic can be seen not only in the time constant but also in the age distribution of PCR test positives. Figure 8 shows the change in the age structure of positive people in Tokyo (Ref. 9), normalized by the number of positive people in their 40s. The age distribution is flat in spring, from February to May in 2020 in Fig.8 (a), and February and March in 2021 shown in Fig.8 (b). In summer, in July and August in 2020 and 2021 shown in Fig.8 (e), the majority of the positives were young people. In the early summer and autumn to winter shown in Figs.8 (c) and (d), the age distributions were between those in spring and summer. From Figure 7 (a) and Figure 8, we can say that the time constant is short when the age distribution is flat. Readers may doubt the seasonality from the data point “W” at the highest positive rate, January 2022. However, this point is another supporting data of the seasonality. We do not know the reason, but the age distribution of the epidemic in January 2022 is that of summer epidemic as seen in Figure 8 (e).

**Fig. 8.**
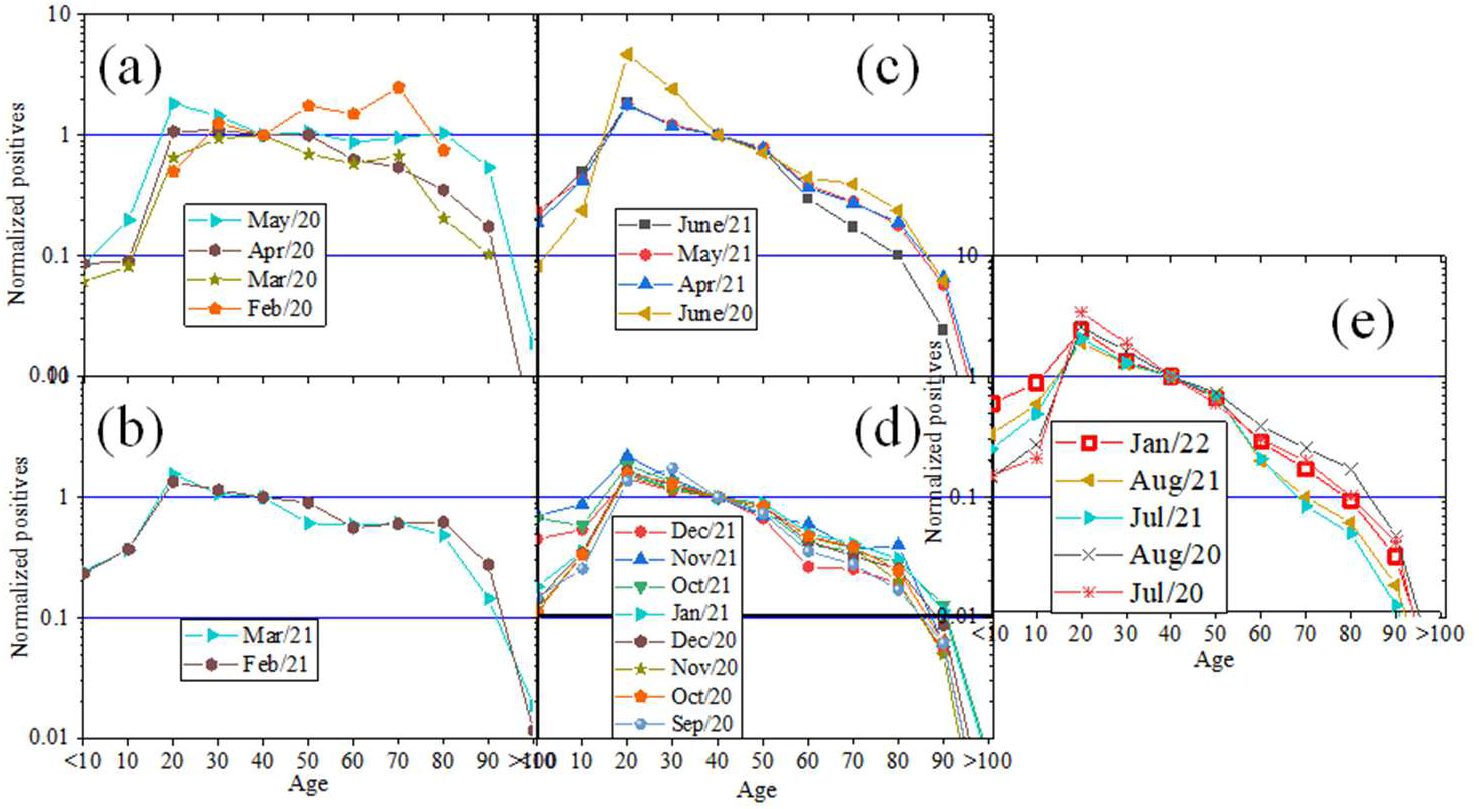
The age distributions of positives in Tokyo normalized by the number of positive people in their 40s. In spring shown in (a) and (b), the distribution was flat. In summer shown in (e), the majority of positives is young people. In transient seasons shown in (c) and (d), the distribution is between those in spring and summer.

The conclusion of Figure 7 (a) and Figure 8 is that the time constant is independent of the peak height but dependent on the age distribution; the epidemic duration is long when most positives are young people.

## 4. Discussions

### 4-1. essential teachings of mathematical models

The essential teachings of the mathematical models are that if *bS*_*0*_ *–g* <0, the spread of the infection does not begin and that the epidemic does not end unless most of the members gain immunity. If the infectivity is high, the number of patients surges, the peak of the epidemic rises, and most people are infected, gaining immunity in a short period. If the infectivity is weak, the epidemic peak is low, and the epidemic lasts for a long time until most people gain immunity. Therefore, the inverse proportionality of the peak height and duration of the epidemics is the essence of mathematical models. The contradiction of Figures 7 (a) and (b) tells that there is a fatal flaw in the current mathematical models. We need to build a new mathematical model for infectious diseases. To get a hint for a new mathematical model, we search for a cause of variation of the peak height of COVID-19.

### 4-2. epidemiological correlation between COVID-19 and rhinovirus

Greer et al. (Ref. 12) reported that the rhinovirus epidemic suppressed the development of other cold viruses. In 660 samples, 53% of collected 1247 samples from 0.5 to 4.3 years old, there were multiple viruses. The number of simultaneous detections with rhinovirus of many viruses such as adenovirus was smaller than expected from the number of single detections of each virus. Wu et al. (Ref. 13) also concluded that rhinovirus suppresses the growth of various viruses from the measurement of simultaneous detection of other viruses. The number of influenza viruses detected was less than one-fifth of the expectation when co-infected with rhinovirus. Linde et al. (Ref. 13) claimed that the increase of rhinovirus prevalence suppressed the influenza pandemic in 2009.

Suggested by these epidemiological studies, we study the correlation between the epidemics of COVID-19 and rhinovirus.

Figure 9 compares the two-year epidemics of COVID-19 and rhinovirus (Ref. 10) in Japan. As indicated by the light blue upward arrows, COVID-19 was prevalent four times at the valleys of the rhinovirus epidemic. As indicated by the purple down-arrows, COVID-19 epidemics were low when a rhinovirus was prevalent.

**Fig. 9.**
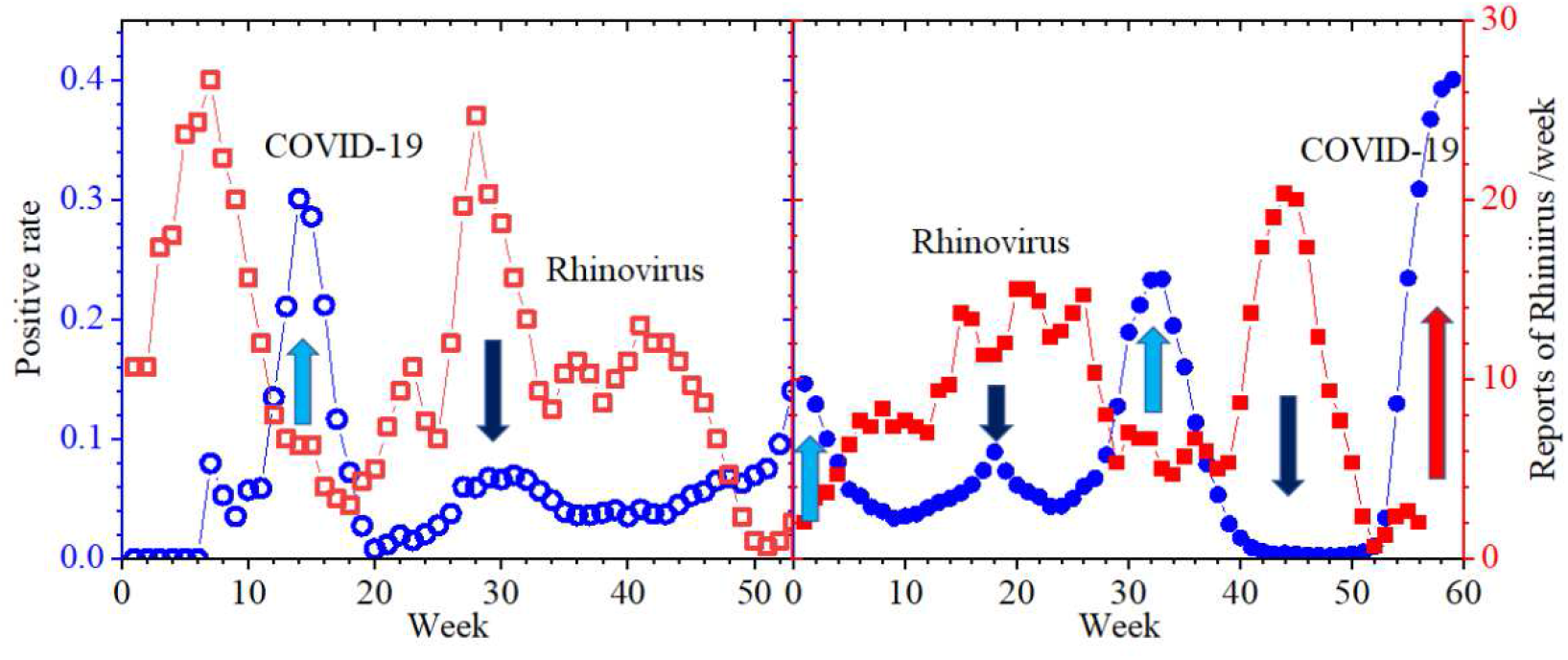
The epidemics of rhinovirus (red squares) and COVID-19 (blue squares) in 2020 and 2021. The COVID-19 and rhinovirus epidemics have reversed profiles.

Figure 10 summarizes the correlation of the epidemics of COVID-19 and rhinovirus. The positive rate at the peak of the COVID-19 epidemics was inversely proportional to the size of the rhinovirus epidemic. The time constants, shown in Fig. 7 (a), are replotted. The COVID-19 epidemic has a good correlation with the prevalence of rhinovirus, although the inverse proportionality with the width of the epidemic, which is the crucial result of the K-M model, is not observed. This fact is a valuable hint for a new model.

**Fig. 10.**
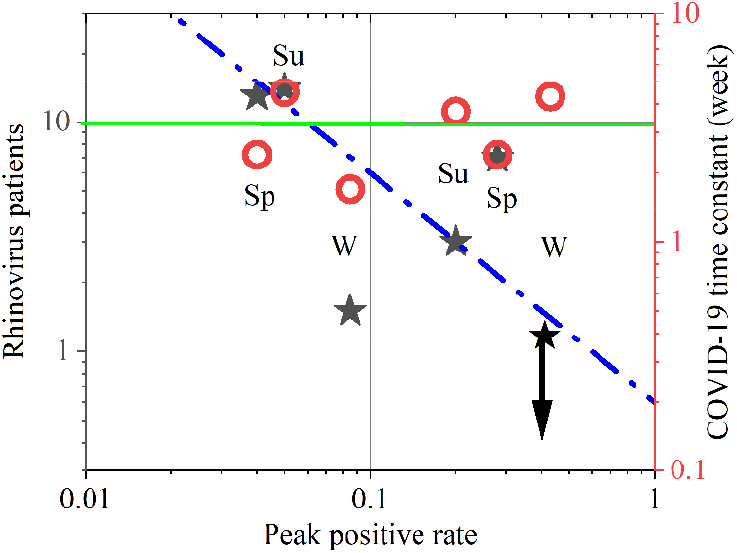
Summary of Fig.8. The positive rate at the peak of the COVID-19 epidemics was inversely proportional to the size of the rhinovirus epidemic. Open red circles are replotting from Fig.7 (a), showing time constants of fitting Gaussians.

The inverse proportionality between the epidemics of COVID-19 and rhinovirus explains the low positive rate in October and November in 2021 seen in Fig.3. Figure 11 shows the COVID-19 and rhinovirus epidemics from the 35th to 53rd weeks in 2020 and 2021. When the positive rate of COVID-19 (filled blue squares) was very low in 2021, there was a high peak of the rhinovirus epidemic (filled red squares). The high prevalence of a rhinovirus suppressed COVID-19. In 2020, the rhinovirus prevalence (open red circles) was a little lower, and the positive rate (open blue circles) in the epidemic valley was not small.

**Fig. 11.**
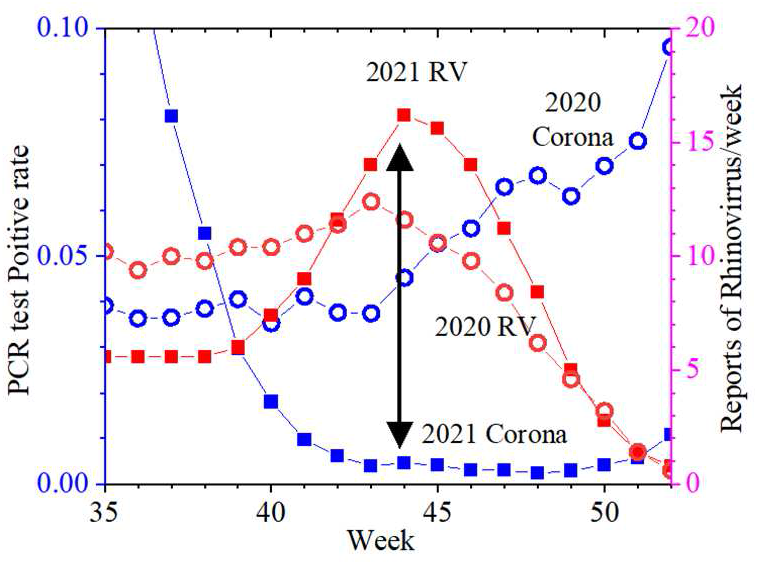
The COVID-19 and rhinovirus epidemics in 2020 and 2021 during the 35th to 52nd weeks, during which the COVID-19 epidemics were in valleys. The positive rate in 2021 was significantly low when the rhinovirus epidemic was large.

The mechanism behind the suppression of the growth of various viruses by the infection of rhinovirus has been clarified in experiments using airway epithelial cells. Wu et al. (Ref. 14) reported the following. When the airway epithelial cells were first infected with rhinovirus and then infected with influenza virus four to five days later, the number of influenza viruses reduced to one-tenth or less. Co-infection with rhinovirus produced much higher antibodies than for the single infection with influenza virus. When BX795, which inhibits the action of interferon, was used in culturing, co-infection with rhinovirus did not reduce the number of influenza viruses, and there was no antibody production. The viral load of rhinovirus did not change regardless of co-infection with influenza virus or the presence or absence of BX795, indicating that the number of antibodies produced by the influenza virus was small. Dee et al. (Ref. 15) showed that rhinovirus infection suppressed the growth of SARS-CoV-2 by generating interferon. The viral load of SARS-CoV-2 increased when infected alone in the epithelial cells of the airways. When rhinovirus was co-infected on the second day, the SARS-CoV-2 decreased rapidly two days later. The addition of SARS-CoV-2 on the second day did not change the number of rhinoviruses, but the viral load of SARS-CoV-2 diminished immediately. When BX795, which inhibits the action of interferon, co-infection with rhinovirus did not block SARS-CoV-2 regeneration. Lei et al. (Ref. 16) showed that interferon suppresses SARS-CoV-2 regeneration.

As described above, the literature has reported that the rhinovirus infection suppresses the growth of SARS-CoV-2 in airway epithelial cells. However, this does not necessarily imply that the prevalence of rhinovirus weakens the COVID-19 epidemic, because the final size, given by 1-exp (-(*R*_*0*_-1)/0.60), increases slowly with the increase of infectious power and interaction between viruses may have a small effect. Hence, our finding of the inverse proportionality between the epidemics of the number of rhinovirus patients will be the first epidemiological confirmation of interference between COVID-19 and rhinovirus. With reports of interferences of rhinovirus with other viruses such as adenovirus and influenza in the literature, we can conclude that rhinovirus has the power of suppressing other viruses.

### 4-3. a novel mathematical model

The variation of the peak height of the COVID-19 epidemic is attributable to the interference with the rhinovirus epidemic. But, the K-M model cannot explain the independence of the duration of the epidemic peak with the height. Equation (3) is approximated as d ln *I(t)*/dt = (*β S*_*0*_ *-γ*) when *S(t)* ≈ *S*_*0*_. For a larger *β*, the increase rate of ln *I(t)* is higher, and the time constant, which is the inverse of the increase rate, is shorter. Thus, the inverse relationship between the peak height and the duration, not observed in actual epidemics, is the inevitable result, implying that the mathematical models have a fatal flaw. By examining Eq. (2), we realize that the fatal flaw originates in the absence of the 0th power term of *I(t)*.

In Eq.2, *S(t)= S*_*0*_ - *I(t)* - *R(t)*, where *R(t)*=∫ *γI* (*t*) *dt*. When the size, *p*(t) =*R(t)/S*_*0*_, is

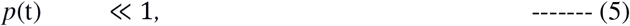

Eq. (2) is approximated as,

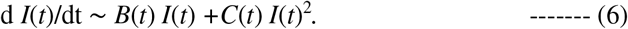

In most actual epidemics, as discussed in the Introduction, the final size *p*(∞) is so small that Eq. (6) is a good approximation for the whole epidemic not only in the early phase.

A general form of a quadratic equation for *I(t)* is

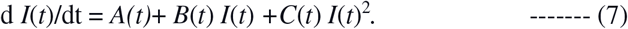

There is no reason for neglecting it, and the term *A*(t) may dominate over other terms in most actual epidemics of infectious diseases. The origin of the critical flaw of the K-M model is the absence of the 0th power term of *I*(t), *A*(t), in Eq. (2).

We expect that the inclusion of the term *A*(t) will also solve two problems mentioned in the Introduction; a far smaller integrated number of patients in actual epidemics than the calculation by mathematical models and constant detection of patients around the year not reproducible by current models.

## 5. Summary

All measures for controlling infectious diseases originate in mathematical models, but there are some problems. We discussed the inverse proportionality between the height and width of the epidemic peak, which is the essential conclusion of mathematical models. Inverse proportionality, however, is not observed in most actual epidemics of infectious diseases.

We examined the inverse proportional relation in the COVID-19 epidemic in Japan. Since spring in 2020, COVID-19 has been prevalent six times in winter, spring, and summer with high seasonality, and the peak size varied by one order of magnitude. The time constants of the six Gaussians that fit the epidemics were independent of peak height, which contradicts the most important conclusion of the mathematical model. We found that the peak height was inversely proportional to the number of rhinovirus patients. The literature has revealed the mechanism behind the discovered force of rhinoviruses in lowering the COVID-19 epidemic.

We discussed that the origin of the flaw of the current models is neglecting the 0th term of infected people in the derivative equation of the K-M model.

## Data Availability

All clinical data analyzed in the present paper are publicly available.
The data of positive people in Tokyo are from the Tokyo Metropolitan Government Corona Virus Infection Control Site. The positive rate is from the file in Monitoring item (4) Positive rate of test. We calculated the age distribution using the data in Attributes of positive people.
The data on rhinovirus is from the file in the Archive of Other Viruses at the site of Infectious Agents Surveillance Report (IASR) at the National Institute of Infectious Disease. 

